# Hospitalizations for emergency-sensitive conditions in Germany during the Covid-19 pandemic Insights from the German-wide Helios hospital network

**DOI:** 10.1101/2021.02.08.21250309

**Authors:** Andreas Bollmann, Sven Hohenstein, Vincent Pellissier, Sebastian König, Laura Ueberham, Gerhard Hindricks, Andreas Meier-Hellmann, Ralf Kuhlen, on behalf of Helios hospitals, Germany

## Abstract

**Background:** While there are numerous reports that describe emergency care during the early Covid-19 pandemic, there is scarcity of data for later stages. This study analyzes hospitalization rates for 37 emergency-sensitive conditions in the largest German-wide hospital network during different pandemic phases.

**Methods:** Using claims data of 80 hospitals, consecutive cases between January 1 and November 17, 2020 were analyzed and compared to a corresponding period in 2019. Incidence-rate ratios (IRR) comparing the both periods were calculated using Poisson regression to model the number of hospitalizations per day.

**Results:** There was a hospitalization deficit between March 12 and June 13, 2020 (coinciding with the 1^st^ pandemic wave) with 32,807 hospitalizations as opposed to 39,379 in 2019 (IRR 0.83, 95% CI 0.82 – 0.85, *P*<0.01). During the following period (June 14 to November 17, 2020, including the start of 2^nd^ wave), hospitalizations were reduced from 63,799 in 2019 to 59,910 in 2020, but this reduction was not that pronounced (IRR 0.94, 95% CI 0.93 – 0.95, *P*<0.01). There was an increase in hospitalizations for acute myocardial infarction, aortic aneurism/dissection and pulmonary embolism after the 1^st^ wave during which hospitalizations had been reduced for those conditions. In contrast, hospitalizations for sepsis, pneumonia, obstructive pulmonary disease, and intracranial injuries were reduced during the entire pandemic.

**Conclusions:** There was an overall reduction of hospitalizations for emergency-sensitive conditions in Germany during the Covid-19 pandemic with heterogeneous effects on different disease categories. The increase of hospitalizations for acute myocardial infarction, aortic aneurism/dissection and pulmonary embolism is an alarming signal that requires attention and further studies.

**KEY MESSAGES:** *What is already known on this subject:*

- There has been a reduction in emergency room visits and hospital admissions for several emergent medical and surgical conditions during the early Covid-19 pandemic (1^st^ wave).

*What this study adds:*

- Using claims data of 80 German-wide Helios hospitals, we found an overall reduction of hospitalizations for emergency-sensitive conditions in Germany during the Covid-19 pandemic until mid November 2020 with heterogeneous effects on different disease categories. While hospitalizations for sepsis, pneumonia, obstructive pulmonary disease, and intracranial injuries were reduced during the entire pandemic. There was an alarming increase of hospitalizations for acute myocardial infarction, aortic aneurism/dissection and pulmonary embolism after the 1^st^ wave.

## INTRODUCTION

While there are numerous reports that describe emergency calls,^1^ emergency room visits ^2^ and hospital admissions ^3-6^ for several medical and surgical conditions during the early Covid-19 pandemic (1^st^ wave), there is scarcity of data during later pandemic stages (e.g. 2^nd^ wave, period between waves). In addition, a comprehensive overview covering previously defined emergency-sensitive conditions ^7^ is also missing.

With this study, we wish to complement previous reports by providing and comparing hospitalization data for patients with emergency-sensitive conditions hospitalized in a large German-wide hospital network during different pandemic phases.

## METHODS

### Study cohort

We performed a retrospective analysis of claims data of 80 Helios hospitals in Germany. Consecutive cases with an emergent hospital admission between January 1 and November 17, 2020 (study period) were analyzed and compared to a corresponding period covering the same weeks in 2019 (control period). Cause-specific hospitalizations were defined based on the primary discharge diagnosis according to International Statistical Classification of Diseases and Related Health Problems [ICD-10-GM (German Modification)] codes for 37 mortality-related emergency-sensitive conditions according to the Panel on Emergency-Sensitive Conditions (Table 1).^7^ Cases with confirmed Covid-19 infection (U07.1) were not excluded from this analysis.

**Table 1.**
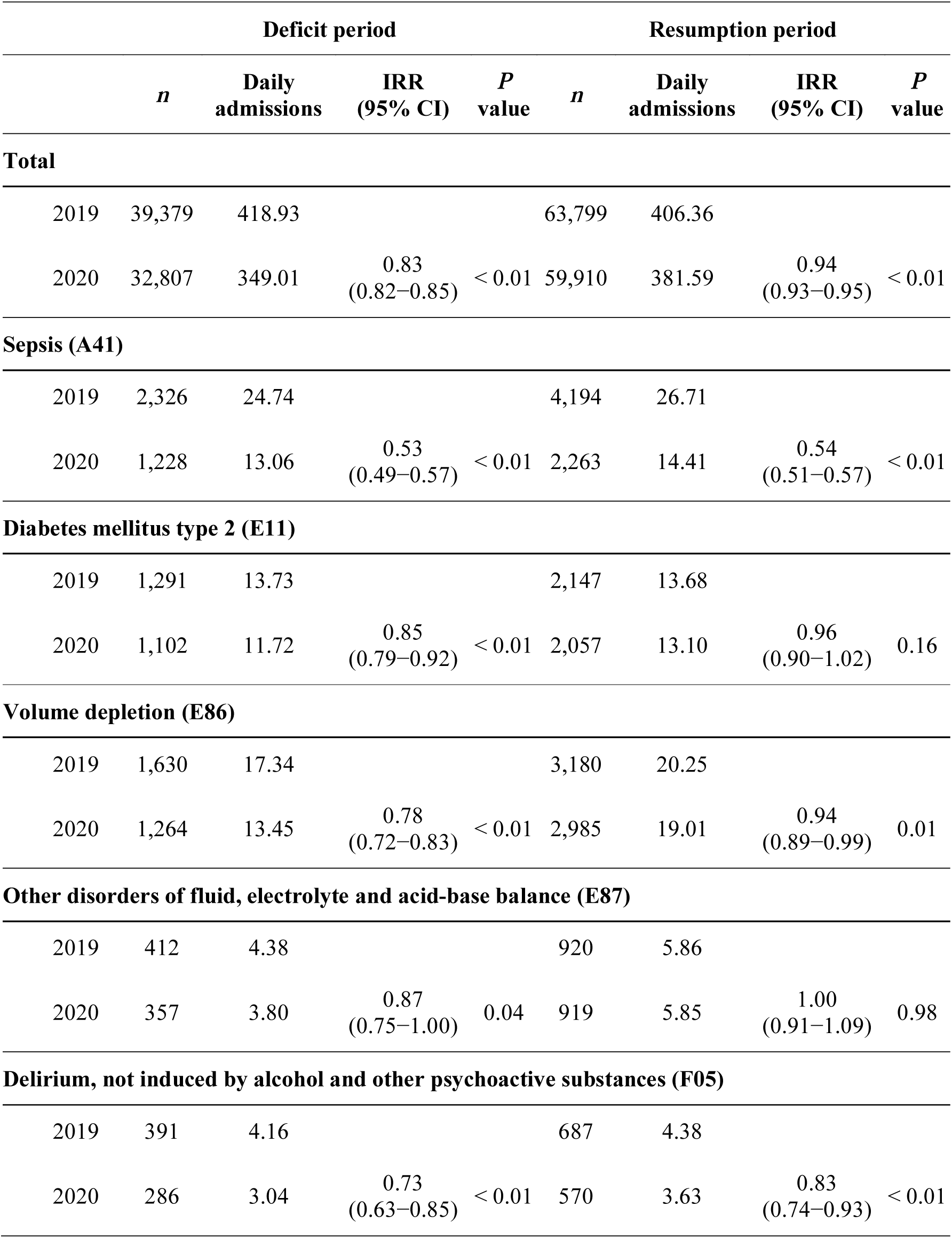
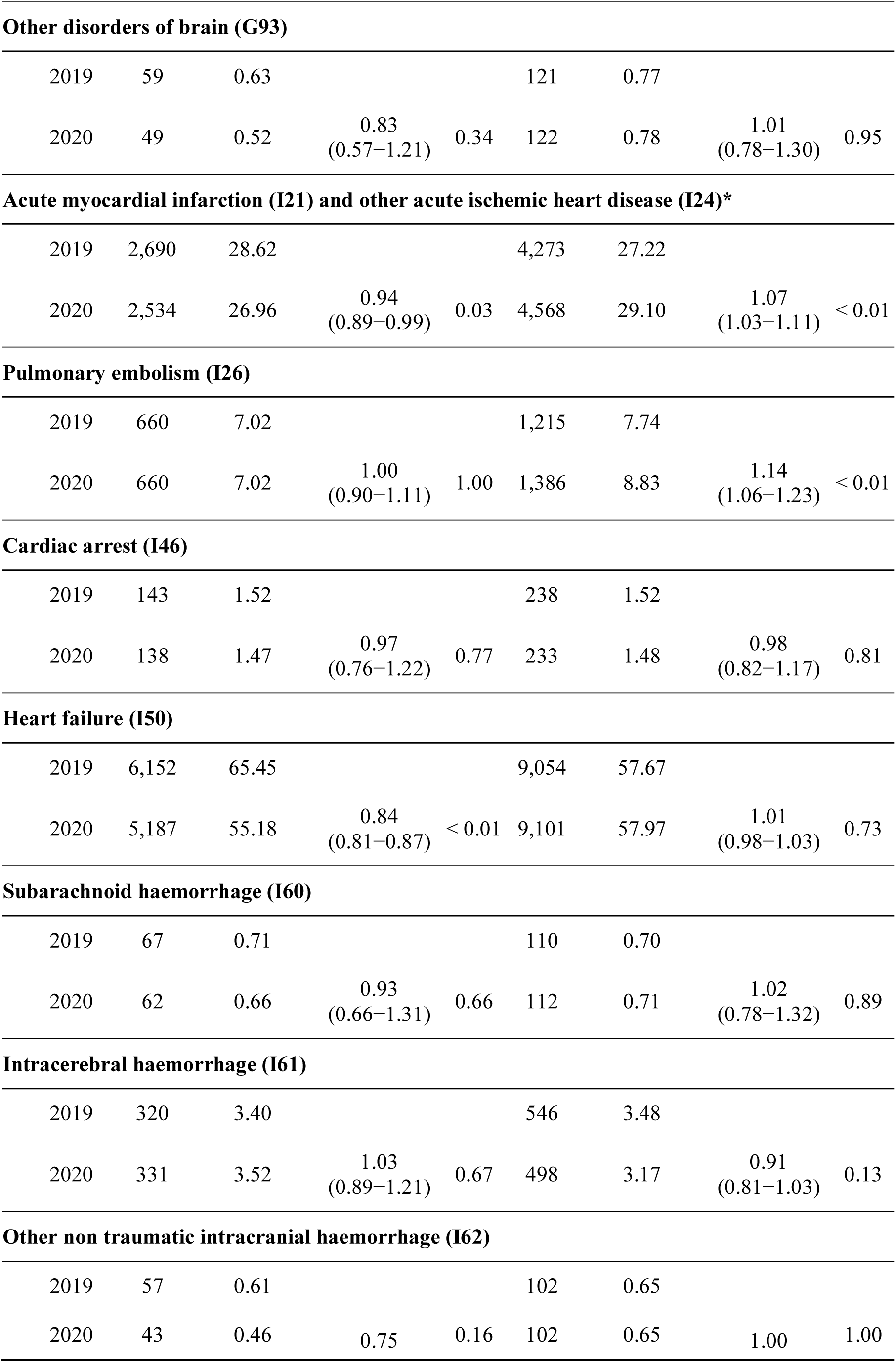
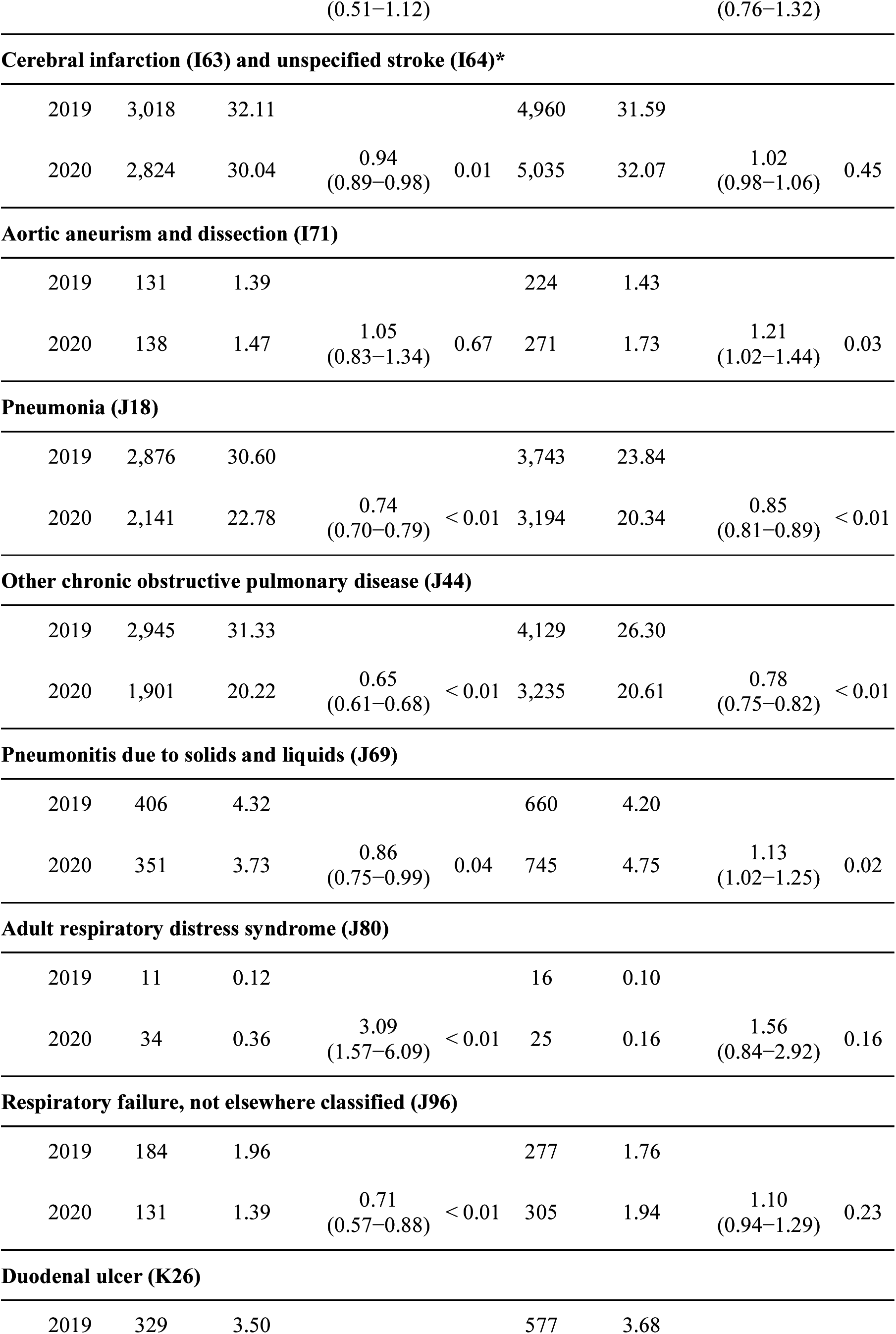
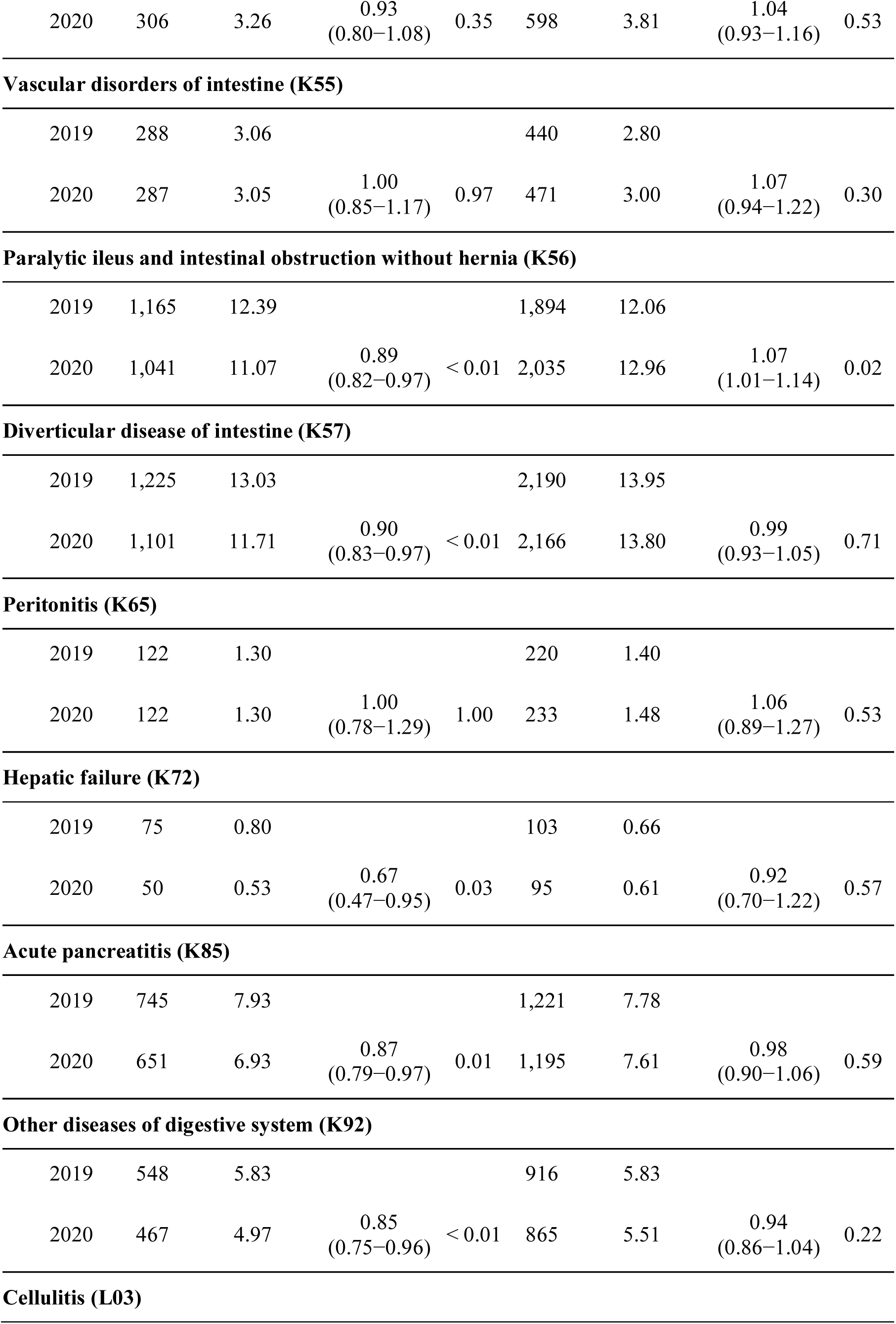
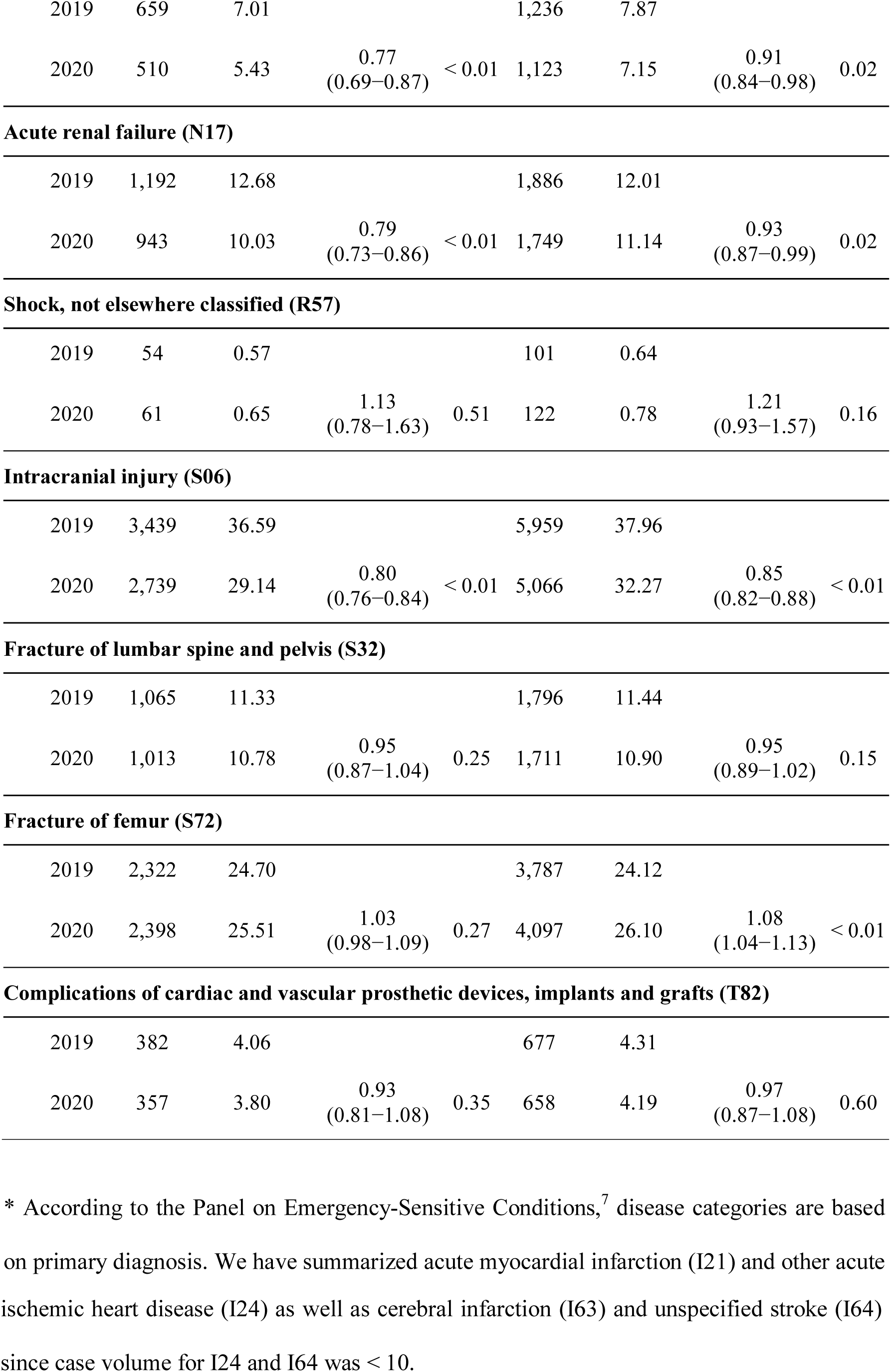
Emergency hospital admissions in the German-wide Helios hospital network during the Covid-19 pandemic. ICD-10 codes for the emergency-sensitive conditions are provided in parenthesis.^7^

This study was approved by the Ethics Committee at the Medical Faculty, Leipzig University (#490/20-ek). Due to the retrospective study of anonymized data informed consent was not obtained. Patients and/or the public were not involved in the design, or conduct, or reporting, or dissemination plans of this research.

### Data Analysis

Administrative data were extracted from QlikView (QlikTech, Radnor, Pennsylvania, USA). Incidence rates for admissions were calculated by dividing the number of cumulative admissions by the number of days for each time period. Incidence-rate ratios (IRR) comparing the study period to the control period were calculated using Poisson regression to model the number of hospitalizations per day. Inferential statistics were based on generalized linear mixed models (GLMM) specifying hospitals as random factor. We report IRR (calculated by exponentiation of the regression coefficients) together with 95% confidence intervals (CI) for the comparisons of the two periods and *P* values for the interactions. For all tests we apply a two-tailed 5% error criterion for significance.

## RESULTS

Hospital admissions for emergency-sensitive conditions and daily new SARS-CoV2 infections in Germany are depicted in Figure 1. There was a hospitalization deficit between March 12 and June 13, 2020 coinciding with the 1^st^ pandemic wave (deficit phase). During this period, there were 32,807 hospitalizations as opposed to 39,379 in 2019 (IRR 0.83, 95% CI 0.82 – 0.85, *P*<0.01) including 286 PCR-confirmed Covid-19 cases. During the following observational period (June 14 to November 17, 2020 including the start of the 2^nd^ infection wave; resumption phase), emergency-sensitive hospitalizations were reduced from 63,799 in 2019 to 59,910 in 2020 including 436 Covid-19 cases, but this reduction was not that pronounced (IRR 0.94, 95% CI 0.93 – 0.95, *P*<0.01) as during the 1^st^ infection wave.

**Figure 1.**
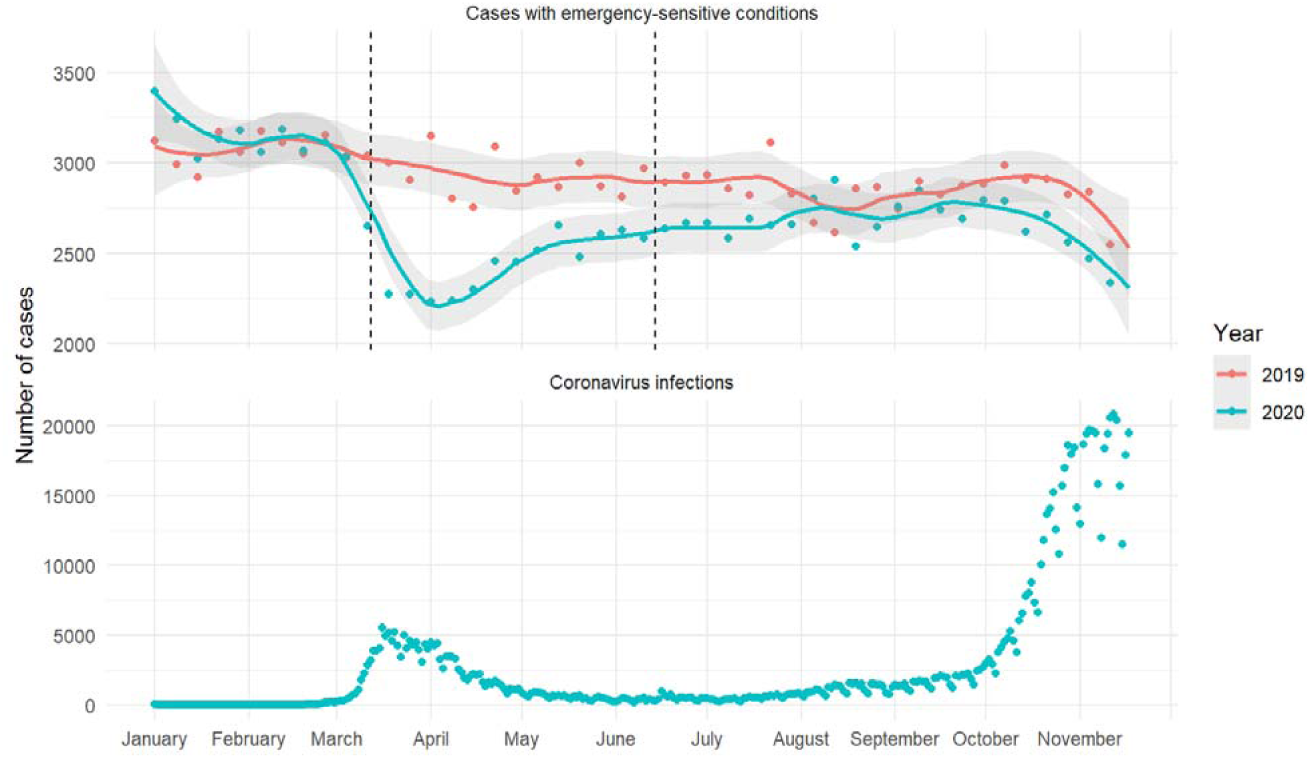
Total weekly hospital admissions for emergency-sensitive conditions (upper panel) and daily new SARS-CoV2 infections in Germany (lower panel). Smooth curves for weekly admission rates were fitted via Locally Weighted Scatterplot Smoothing (LOESS) with a degree of smoothing of α = 0.25. Grey areas represent 95% confidence intervals (CI). A 2020 deficit period starting at the first the day with non-overlapping 95% CI intervals and ending at the last day with non-overlapping intervals was defined. The first day at which the intervals are overlapping again, was defined as the start of the resumption period (dotted vertical lines).

Hospitalizations for the individual emergency-sensitive conditions stratified for both periods are summarized in Table 1 showing a heterogeneous pattern. Of note, despite the inclusion of Covid-19 cases, hospitalizations with sepsis, pneumonia, obstructive pulmonary disease, and intracranial injuries as primary diagnosis were reduced during the entire pandemic. Similarly, when expanding the definition of pneumonia (J18) to include all severe acute respiratory infections (J09 – J22, excluding hospital acquired infections U69.0), IRR was 0.76 (95% CI 0.74 – 0.78, *P*< 0.01) in the deficit and 0.97 (95% CI 0.94 – 0.99, *P*< 0.01) in the resumption phase. In contrast, there was an increase in hospitalizations for acute myocardial infarction (IRR 1.07, 95% CI 1.03 –1.11, *P* < 0.01), aortic aneurism and dissection (IRR 1.21, 95% CI 1.02 – 1.44), *P*=0.03) and pulmonary embolism (IRR 1.14, 95% CI 1.06 – 1.23, *P* < 0.01) after the 1^st^ infection wave.

## DISCUSSION

This study was performed in the largest German-wide hospital network serving about 10% of the German population by analyzing emergency-sensitive conditions from claims data. This comprehensive list ^7^ has been suggested for the assessment of the acute care system but modifications (e.g. expansion of definitions for unspecific pneumonia, J18) may be warranted.^8^

We found an initial reduction in several emergency-sensitive conditions including myocardial infarction, heart failure, diabetes mellitus, or pancreatitis that is in agreement with previous studies.^2-6^ Those observations corresponded with the initiation of national public health emergency measures, and this decline persisted for several weeks. On the one hand, there may be a true reduction in the incidence of emergencies as a result of lower physical or psychological stress, improved medication adherence, diminished air pollution, traffic, and infectious disease transmission, or better outpatient care delivery models. In fact, the consistently lower rates of exacerbations of respiratory conditions, or traumatic intracranial injuries support this hypothesis. On the other hand, it is the possibility that patients were reluctant to seek medical attention due to fear of contagion at the hospital. In addition, the emphasis on social distancing might have inappropriately persuaded patients to avoid in-person medical care. While this reduction was pronounced during the 1^st^ wave and effected multiple conditions, hospitalizations for the majority of those resumed to previous year levels during later pandemic phases.

Of special concern is the increased incidence of hospitalizations for acute myocardial infarction, aortic aneurism and dissection as well as pulmonary embolism. While the former may be a result of reduced cardiovascular care during the early pandemic,^5,6^ the latter could also be associated with preceding Covid-19 infections.^9^ If the increased incidence of hospitalizations for acute myocardial infarction, aortic dissection and pulmonary embolism is a signal for a rising incidence of those conditions in the public, this could at least in part explain the observed excess mortality in Germany between late July and mid October 2020.^10^ Those alarming signals require immediate attention and further studies.

## Data Availability

The data that support the findings of this study are available on request from the corresponding author.

## REFERENCES

1. Valent F, Licata S. Emergency medical services calls during Italy’s COVID-19 lockdown. Ann Emerg Med. 2020 Dec;76(6):812–814. doi:10.1016/j.annemergmed.2020.06.036.

2. Westgard BC, Morgan MW, Vazquez-Benitez G, Erickson LO, Zwank MD. An analysis of changes in emergency department visits after a state declaration during the time of COVID-19. Ann Emerg Med. 2020 Nov;76(5):595–601. doi:10.1016/j.annemergmed.2020.06.019.

3. Anderson TS, Stevens JP, Pinheiro A, Li S, Herzig SJ. Hospitalizations for emergent medical, surgical, and obstetric conditions in Boston during the COVID-19 pandemic. J Gen Intern Med. 2020 Jul 22:1–4. doi:10.1007/s11606-020-06027-2.

4. Baum A, Schwartz MD. Admissions to Veterans Affairs hospitals for emergency conditions during the COVID-19 pandemic. JAMA. 2020 Jul 7;324(1):96–99. doi:10.1001/jama.2020.9972.

5. Bollmann A, Hohenstein S, Meier-Hellmann A, Kuhlen R, Hindricks G. Emergency hospital admissions and interventional treatments for heart failure and cardiac arrhythmias in Germany during the Covid-19 outbreak: insights from the German-wide Helios hospital network. Eur Heart J Qual Care Clin Outcomes. 2020 Jul 1;6(3):221–222. doi:10.1093/ehjqcco/qcaa049.

6. Bollmann A, Pellissier V, Hohenstein S, König S, Ueberham L, Meier-Hellmann A, Kuhlen R, Thiele H, Hindricks G; Helios hospitals, Germany. Cumulative hospitalization deficit for cardiovascular disorders in Germany during the Covid-19 pandemic. Eur Heart J Qual Care Clin Outcomes. 2020 Aug 28:qcaa071. doi:10.1093/ehjqcco/qcaa071.

7. Berthelot S, Lang ES, Quan H, Stelfox HT; Panel on Emergency-Sensitive Conditions (PESC). Identifying emergency-sensitive conditions for the calculation of an emergency care inhospital standardized mortality ratio. Ann Emerg Med. 2014 Apr;63(4):418-24.e2. doi:10.1016/j.annemergmed.2013.09.016.

8. Vashi AA, Urech T, Carr B, Greene L, Warsavage T Jr, Hsia R, Asch SM. Identification of emergency care-sensitive conditions and characteristics of emergency department utilization. JAMA Netw Open. 2019 Aug 2;2(8):e198642. doi:10.1001/jamanetworkopen.2019.8642.

9. Vlachou M, Drebes A, Candilio L, Weeraman D, Mir N, Murch N, Davies N, Coghlan JG. Pulmonary thrombosis in Covid-19: before, during and after hospital admission. J Thromb Thrombolysis. 2021 Jan 1. doi: 10.1007/s11239-020-02370-7.

10. König S, Hohenstein S, Ueberham L, Hindricks G, Meier-Hellmann A, Kuhlen R, Bollmann A. Regional and temporal disparities of excess all-cause mortality for Germany in 2020: Is there more than just COVID-19? J Infect. 2020 Dec 23:S0163-4453(20)30777-5. doi: 10.1016/j.jinf.2020.12.018.

